# Evaluation of the auxiliary diagnosis value of antibodies assays for the detection of novel coronavirus (SARS-Cov-2)

**DOI:** 10.1101/2020.03.26.20042044

**Authors:** Gao Yong, Yuan Yi, Li Tuantuan, Wang Xiaowu, Li Xiuyong, Li Ang, Han Mingfeng

## Abstract

**Background:** The spread of an novel coronavirus (SARS-CoV-2, previously named 2019-nCoV) has already taken on pandemic proportions, affecting over 100 countries in a matter of weeks. Elucidating the diagnostic value of different methods, especially the auxiliary diagnosis value of antibodies assays for SARS-CoV-2 infection is helpful for improving the sensitivities of pathogenic-diagnosis, providing timely treatment, and differentiating the infected cases from the healthy, thus preventing further epidemics.

**Methods:** Medical records from 38 patients with confirmed SARS-CoV-2 infection in the Second People’s Hospital of Fuyang from January 22, 2020 to February 28, 2020 were collected and retrospectively analyzed. Specimens including throat swabs, sputum and serum were collected during the hospitalization period, viral RNAs and serum IgM-IgG antibodies to SARS-CoV-2 were measured respectively. The detectability of different methods as well as the auxiliary diagnosis value of antibodies test for SARS-CoV-2 infection were analyzed.

**Results:** Among 38 patients, the total seropositive rate for IgM and IgG was 50.0% and 92.1%, respectively. Two patients remained seronegative throughout the course of illness. In the early phase of illness, the RNA test for sputum specimens possessed the highest detectability(92.3%), followed by the the RNA test for throat swabs (69.2%), and the antibodies assays presented lower positive rates(IgM, 23.0%, IgG, 53.8%). While, the sensitivity of antibodies assays overtook that of RNA test since day 8 after onset (IgM, 50.0%; IgG, 87.5%). Of note, the positive rate of throat swabs was only 13.0% for cases in later phase(≥15 d.a.o), and the sensitivities of IgM and IgG rose to 52.2% and 91.3%, respectively. Combined use of antibodies assay and qRT-PCR at the same time was able to improve the sensitivities of pathogenic-diagnosis, especially for the throat swabs group at the later stage of illness. Moreover, most of these cases with undetectable viral RNA in throat swabs specimens at the early stage of illness were able to be IgM/IgG seropositive after 7 days.

**Conclusions:** The antibodies detection against SARS-CoV-2 offers vital clinical information for physicians, and could be used as an effective supplementary indicator for suspected cases of negative viral nucleic acid detection or in conjunction with nucleic acid detection in the diagnosis of suspected cases.

## Introduction

The outbreak of coronavirus disease 2019 (COVID-19), caused by the severe acute respiratory syndrome coronavirus 2 (SARS-CoV-2) infection, continues to spread worldwide and has become a global threat. As of March 21, 2020, it has reached more than 115 countries, with 266073 cases and 11184 deaths(1-2). The World Health Organization has declared the COVID-19 outbreak a pandemic and rates the global risk assessment as very high.A global response to prepare health systems worldwide is imperative(3).Timely and accurate diagnosis of suspected SARS-CoV-2 infection, and to isolate and care for these patients early are of great importance for interrupting human-to-human transmission, and limiting further spread of the virus. Common laboratory abnormalities in confirmed COVID-19 cases include decreased white blood cells, lymphocytes, and platelet counts, and an increased LDH, CK, and CRP levels(4-5). And the reported common clinical symptoms include fever, cough, myalgia or fatigue(6). However, these abnormalities and symptoms are not unique features of COVID-19 since these might be similar to that of other virus infected disease. Meanwhile, some infected patients are asymptomatic but can also become a source of infection, which makes early diagnosis essential.

The use of quantitative real-time PCR (qRT-PCR) assays for the detection of the viral nucleic acid has become the primary and crucial diagnostic approach for identification of SARS-CoV-2 infection, while it still has some limitations in clinical practice(7-9). The RNA-based diagnostic tests only give a positive result when the virus is still present. The tests could not identify people who has been infected in the past, recovered, and cleared the virus from their bodies. In addition, negligible false-negative risk brought by PCR test were reported and the positive rates varied for different specimens in COVID-19 patients. A number of cases that were epidemiologically linked to SARS-CoV-2 exposure and with typical lung CT images still remained RNA negative in their respiratory tract specimens(10-12).

IgG/IgM antibodies test, a serological test method, has been added as a diagnostic criteria in China’s updated version of the diagnosis and treatment guidelines for COVID-19 (3rd, March).It will help to trace in a much more population-based way whether a person has been infected in the past since the body could retain antibodies against virus that it has already overcome.While such assays still need to be carefully validated to be sure they could react only to antibodies against SARS-CoV-2. The similarity between SARS-CoV-2 and the other viruses might lead to cross-reactivity. Also here were still false positive and false negative results for IgG/IgM antibodies test(13-15). Herein, there is an urgent need for elucidating the diagnostic accuracy of different specimens and methods to improve the positive rates, so as to prevent virus transmission and to assure timely treatment of patients.

## Materials and Methods

### Study Design and Participants

Medical records from 38 COVID-19 patients (aged from 15 years to 75 years) in the Second People’s Hospital of Fuyang from January 22, 2020 to February 28, 2020 were collected and retrospectively analyzed. Diagnosis of COVID-19 was based on the New Coronavirus Pneumonia Prevention and Control Program (5th edition) published by the National Health Commission of China. Specimens including throat swabs, sputum and serum were collected during the hospitalization period.Viral RNAs and serum IgM-IgG antibodies against SARS-CoV-2 were measured by qRT-PCR (Real-Time Reverse Transcription Polymerase Chain Reaction Assay) and GICA assay (Colloidal Gold Antibodies Test), respectively. This study was approved by the National Health Commission of China and Ethics Commission of the Second People’s Hospital of Fuyang. Written informed consent was waived by the Ethics Commission of the designated hospital for emerging infectious disease and the urgent need to collect data.

### qRT-PCR Assay for SARS-CoV-2

Respiratory specimens including throat swabs and sputum were collected, and then the throat swabs were placed into a sterile test tube with 1 mL sterile saline, the sputum samples were added equal volume of acetylcysteine and shaken at room temperature for 30 min to be fully liquefied. Next, total RNA was extracted using a viral nucleic acid isolation kit (Jiangsu Bioperfectus Technologies Company, Ltd.), and the qRT-PCR assay was performed using a SARS-CoV-2 nucleic acid detection kit according to the manufacturer’s instructions (Shanghai BioGerm Medical Biotechnology Co.,Ltd.). The open reading frame 1ab (ORF1ab) and nucleocapsid protein (N) genes of SARS-CoV-2, were simultaneously amplified and tested. (1) Interpretation of test results: ① FAM channel for ORFlab gene detection, HEX/C Channel for N gene detection; ② Negative results: Ct value> 37 or not detected; ③ Positive results: amplification curve was S-shaped, and Ct value ≤ 37;④ Suspicious results: the amplification curve was S-shaped, and 37 <Ct value <40. (2) Criteria for SARS-CoV-2-infection interpretation: ① Both of the two genes (ORFla / b, N) of SARS-CoV-2 in one specimen were positive; ② Cases with a single positive gene required confirmation by retesting. If it is still positive for the same single target, it is determined to be positive.If not, it is determined to be negative.These diagnostic criteria were based on the recommendation by the National Institute for Viral Disease Control and Prevention of China (http://ivdc.chinacdc.cn/kyjz/202001/t20200121_211337.html).

### Colloidal Gold Antibodies Test for SARS-CoV-2

Serum IgG and IgM antibodies against SARS-CoV-2 were tested by using a GICA kits according to the manufacturer’s protocol (Innovita Biological Technology Co., Ltd.). Briefly, for each test, 10μL of serum sample and 80μL of sample diluent were added onto the pad of the test strip. Then the strip was placed flat at room temperature for 15min to react and then result could be judged according to the color of the tested and control lines. ① Both the detection band and the control band turn red, the sample will be interpreted as positive.② If the control band turns red while the detection band does not, it will be interpreted as negative. ③ If neither band was colored, the test reagents will be assumed to be not working and required confirmation by retesting.

### Statistical Analysis

All analyses were performed using SPSS 19.0. Continuous variable data were in the median (Interquartile range, IQR), categorical variables were expressed as frequencies (percentages), chi-square test with Yates’s correction or Fisher’s exact test was used for comparison between groups. *P*<0.05 was considered to be statistically significant.

## Results

### Clinical characteristics and seropositive rates of antibodies against SARS-CoV-2

Medical records from 38 COVID-19 patients were collected and retrospectively analyzed, the median age was 40.5 years (IQR, 31.0-49.5years) and 55.3% were males. Of these patients, 3 cases were in severe or critical conditions, and the rest were mild cases. The median number of specimens collected from each patient was 8. A total of 76 serum samples collected during the hospitalization period of the above 38 patients were tested for IgM and IgG antibodies against SARS-CoV-2. The total seropositive rate for IgM and IgG was 50.0%(19/38)and 92.1%(35/38), respectively. Two patients remained seronegative for antibodies testing during hospitalization.Both of the them are close contacts. Case 1 was a female aged 15 with no fever or fatigue, no symptoms of digestive system, and no significant change in lymphocyte subsets counts during the course of disease. The antibodies test still showed to be seronegative in the 14 days following hospital discharge. Case 2 was a female aged 40 with a fever, body temperature up to 38.2°C in the onset of illness. CT scan of chest showed a sign of inflammation, accompanied by an increased T-lymphocyte subsets counts and a decreased NK cells counts. The antibodies test showed a seroconversion of IgG in the 14 days following hospital discharge.

### The detectability of RNA test and antibodies assays for patients in different time after onset

We analyzed the detectability of RNA test and antibodies assays according to the time course since illness onset in the cohort. As the results shown in **Table 1**, in the early phase of illness within 7-day since onset, the RNA test for sputum specimens possessed the highest detectability of 92.3%, followed by the the RNA test for throat swabs (69.2%), and the antibodies assays presented lower positive rates(IgM, 23.0%; IgG, 53.8%). While, the sensitivity of antibodies assays overtook that of RNA test since day 8 after onset (IgM, 50.0%; IgG, 87.5%). Of note, the positive rate of throat swabs was only 13.0% for cases in later phase(≥15 d.a.o), and the sensitivities of IgM and IgG rose to 52.2% and 91.3%, respectively.

**Table 1.** Different detections in samples at different time since onset of patients.

| d.a.o | n | RNA for sputum |  | RNA for throat swabs |  | IgM |  | IgG |  |
| --- | --- | --- | --- | --- | --- | --- | --- | --- | --- |
|  |  | n<sup>+</sup> | Sensitivity | n<sup>+</sup> | Sensitivity | n<sup>+</sup> | Sensitivity<br>y | n<sup>+</sup> | Sensitivity |
| Total | 38 | 29 | 76.3% | 14 | 36.8% | 19 | 50.0% | 35 | 92.1% |
| 0-7 | 13 | 12 | 92.3% | 9 | 69.2% | 3 | 23.0% | 7 | 53.8% |
| 8-14 | 8 | 3 | 37.5% | 2 | 25.0% | 4 | 50.0% | 7 | 87.5% |
| ≥15 | 23 | 14 | 60.8% | 3 | 13.0% | 12 | 52.2% | 21 | 91.3% |

### The auxiliary diagnosis value of antibodies assays for suspected cases of negative nucleic acid detection

Based on the above mentioned findings, we aimed to evaluate the auxiliary diagnosis value of antibodies assays for suspected cases of negative nucleic acid detection. Firstly, the detectability of antibodies assays in patients with undetectable viral RNA in their respiratory tract specimens were analyzed. As the results shown in **Table 2** and **Table 3**, combined use of the tests of RNA and antibodies assay at the same time was able to improve the sensitivities of pathogenic-diagnosis for SARS-CoV-2 infection, especially for the throat swabs group at the later stage of illness.Then, we further analyzed the antibodies test data of cases with undetectable viral RNA in their throat swabs specimens at the early stage of illness. Most of them were shown to be IgM/IgG seropositive after 7 days of negative nucleic acid test results (IgM^+^ 47.1%, IgG^+^ 91.1%), suggesting an auxiliary diagnosis value of antibodies assays.

**Table 2.** **Presence of antibodies against SARS-CoV-2 in cases with undetectable viral nucleic acid in their sputum specimens**

| d.a.o | No of cases with undetectable RNA | IgM |  | IgG |  |
| --- | --- | --- | --- | --- | --- |
|  |  | n<sup>+</sup> | Sensitivity | n<sup>+</sup> | Sensitivity |
| 0-7 | 1 | 0 | 0.0% | 0 | 0.0% |
| 8-14 | 5 | 3 | 60.0% | 5 | 100.0% |
| ≥15 | 9 | 4 | 44.4% | 9 | 100.0% |

**Table 3.** **Presence of antibodies against SARS-CoV-2 in cases with undetectable viral nucleic acid in their throat swabs specimens**

| d.a.o | No of cases with undetectable RNA | IgM |  | IgG |  |
| --- | --- | --- | --- | --- | --- |
|  |  | n<sup>+</sup> | Sensitivity | n<sup>+</sup> | Sensitivity |
| 0-7 | 4 | 2 | 50.0% | 4 | 100.0% |
| 8-14 | 6 | 2 | 33.3% | 6 | 100.0% |
| ≥15 | 20 | 9 | 45.0% | 18 | 90.0% |

## Discussion

The spread of a novel coronavirus (SARS-CoV-2) has recently been identified in patients with acute respiratory disease.It is the third highly pathogenic and transmissible coronavirus after severe acute respiratory syndrome coronavirus (SARS-CoV) and Middle East respiratory syndrome coronavirus (MERS-CoV) emerged in humans(16). Effective diagnosis that identify SARS-CoV-2 infection in people are of great importance for public health efforts, not just for individuals’ health concerns. Patients’ clinical manifestations mainly included fever, cough, dyspnea, myalgia, fatigue, and radiographic evidence of pneumonia. Diagnosis could be based on clinical history, laboratory and CT images, but confirmation primarily relies on nucleic acid detection. Yet many novel coronavirus pneumonia patients can not be diagnosed due to negative nucleic acid test. For example, throat swab is commonly used for nucleic acid test, while the viral loads in upper respiratory tract samples are usually much lower than that in lower respiratory tract samples in COVID-19 cases, and the viral loads of patients varies in different stage of illness(17).

Consequently, these problems lead to an urgent need for clinical auxiliary diagnosis, so as to improve the positive detective rates and to provide timely treatment and preventive quarantine. The human immune system can produce specific IgM and IgG antibodies against virus infection. IgM is the earliest antibody that appears upon first immune response. Serological presence of IgM indicates a recent infection and could be used as auxiliary diagonosis of early infection. IgG is produced later and lasts long, which can be used as an indicator of previous or secondary infection(18-19). In this study, the positive rate of throat swabs was only 13.0% for cases in later phase(≥ 15 d.a.o), and the sensitivities of IgM and IgG were 52.2% and 91.3%, respectively. Combining use of antibodies assay at the same time could be able to greatly improve the sensitivities of pathogenic-diagnosis for SARS-CoV-2 infection, especially for the throat swabs group during the later stage of illness.Moreover, through further analysis of the antibodies test data of cases with undetectable viral RNA in throat swabs specimens at the early stage of illness, we suggest that most of these cases could be IgM/IgG seropositive after 7 days of negative nucleic acid test results, indicating an auxiliary diagnosis value of antibodies assays.

It should be noted that there were still false positive and false negative results for antibodies assay. When IgM and IgG levels are below the detection limit, the test results would be negative. Furthermore, IgM antibodies would gradually decrease and disappear after 2 weeks, so that the IgM level might be below its peak and not detectable in some cases.Meanwhile, the difference in individual immune response might result in the false negative results in suspected cases(19-20). Patients who do not produce antibodies, or who produce antibodies relatively late, might have a relapse once their immunity is reduced.

In conclusion, the test of IgM and IgG antibodies against SARS-CoV-2 provides important immunological evidence for physicians, and could be used as an effective supplementary indicator for suspected cases of negative viral nucleic acid detection or in conjunction with nucleic acid detection in the diagnosis of suspected cases.

Combination of nucleic acid RT-PCR and IgM-IgG antibodies test can provide more accurate SARS-CoV-2 infection diagnosis.More research and development of the SARS-CoV-2 IgG-IgM combined antibodies test kit need to be encouraged to improve the diagnostic sensitivity and specificity for patients.

## Data Availability

The corresponding authors had full access to all data in the study and take responsibility for the integrity of the data and the accuracy of the data analysis.

## Authors’ contributions

Gao Yong, Yuan Yi, Wang Xiaowu, Li Ang and Han Mingfeng were involved in designing the study and manuscript preparation; Gao Yong and Li Tuantuan performed most of experiments;Gao Yong, Yuan Yi, Li Tuantuan and Wang Xiaowu analyzed the data; Gao Yong,Yuan Yi, Li Ang, Li Xiuyong and Han Mingfeng contributed to critical revision of the report. The corresponding authors were responsible for all aspects of the study to ensure that issues related to the accuracy or integrity of any part of the work were properly investigated and resolved. All authors reviewed and approved the final version of the manuscript.

